# Evaluation of serological tests for SARS-CoV-2: Implications for serology testing in a low-prevalence setting

**DOI:** 10.1101/2020.05.31.20118273

**Authors:** Katherine Bond, Suellen Nicholson, Seok Ming Lim, Theo Karapanagiotidis, Eloise Williams, Douglas Johnson, Tuyet Hoang, Cheryll Sia, Damian Purcell, Sharon R Lewin, Mike Catton, Benjamin P Howden, Deborah A Williamson

**Affiliations:** Department of Microbiology, Royal Melbourne Hospital, Melbourne, Australia; Department of Microbiology and Immunology, The University of Melbourne at The Peter Doherty Institute for Infection and Immunity, Melbourne, Australia; Victorian Infectious Diseases Reference Laboratory at The Peter Doherty Institute for Infection and Immunity, Melbourne, Australia; Department of General Medicine and Infectious Diseases Royal Melbourne Hospital, Melbourne, Australia; Departments of General Medicine, The University of Melbourne, Australia; Microbiological Diagnostic Unit Public Health Laboratory, Department of Microbiology and Immunology, The University of Melbourne at The Peter Doherty Institute for Infection and Immunity, Melbourne, Australia; The Peter Doherty Institute for Infection and Immunity, Royal Melbourne Hospital and The University of Melbourne, Melbourne, Australia; Department of Infectious Diseases, Alfred Hospital and Monash University, Melbourne, Australia

**Keywords:** COVID-19, serology, lateral flow, ELISA, neutralisation

## Abstract

**Background:** Robust serological assays are essential for long-term control of the COVID-19 pandemic. Many recently released point-of-care (PoCT) serological assays have been distributed with little pre-market validation.

**Methods:** Performance characteristics for five PoCT lateral flow devices approved for use in Australia were compared to a commercial enzyme immunoassay (ELISA) and a recently described novel surrogate virus neutralisation test (sVNT).

**Results:** Sensitivities for PoCT ranged from 51.8% (95% CI 43.1 to 60.4%) to 67.9% (95% CI 59.4–75.6%), and specificities from 95.6% (95% CI 89.2–98.8%) to 100.0% (95% CI 96.1–100.0%). Overall ELISA sensitivity for either IgA or IgG detection was 67.9% (95% CI 59.4–75.6), increasing to 93.8% (95% CI 85.0–98.3%) for samples > 14 days post symptom onset. Overall, sVNT sensitivity was 60.9% (95% CI 53.2–68.4%), rising to 91.2%% (95% CI 81.8–96.7%) for samples collected > 14 days post-symptom onset, with a specificity 94.4% (95% CI 89.2–97.5%),

**Conclusion:** Performance characteristics for COVID-19 serological assays were generally lower than those reported by manufacturers. Timing of specimen collection relative to onset of illness or infection is crucial in the reporting of performance characteristics for COVID-19 serological assays. The optimal algorithm for implementing serological testing for COVID-19 remains to be determined, particularly in low-prevalence settings.

## INTRODUCTION

The coronavirus disease (COVID-19) pandemic caused by severe acute respiratory syndrome coronavirus 2 (SARS-CoV-2) is a global public health emergency on an unprecedented scale. First reports in late December 2019 described a cluster of patients with pneumonia, linked to a live animal market in Wuhan, China (1–3). One of the fundamental pillars in the prevention and control of COVID-19 is timely, scalable and accurate diagnostic testing. Diagnostic testing plays a critical role in defining the epidemiology of the disease, informing case and contact management, and ultimately in reducing viral transmission (4). To date, testing has comprised detection of SARS-CoV-2 virus using reverse-transcriptase PCR (RT-PCR) assays, predominantly from patients meeting specific epidemiological criteria. However, the immense scale of RT-PCR diagnostic testing has placed extraordinary demands on laboratories, with challenges relating to supply chains of swabs, laboratory reagents, and the human and financial resource required to support population-level testing.

Over the past two months, there has been rapid development of serological assays for COVID-19 in a number of countries (5). Serological tests rely on detection of specific anti-viral antibodies (IgM, IgG, IgA or total antibody) in patient serum, plasma or whole blood. The broad array of serological tests now available vary both in analytical performance and in their particular utility in the overall public health response to COVID-19. The most publicised serological tests for COVID-19 have been lateral flow immunoassays, also known as serological point of care tests (PoCT), which have been manufactured and deployed in several countries. Most available PoCT involve detection of anti-SARS-CoV-2 IgM or IgG antibodies through binding to immobilised antigen (generally domains of the spike (S) protein) attached to colloidal gold, followed by detection of the conjugates by an anti-human IgM or IgG antibody. In addition, a control line is usually also included in the assay, which helps determine whether the test result is valid. The relatively cheap and simple nature of lateral flow assays means that production is suited to scaling-up for increased testing capacity.

In many countries, including the United States and Australia, the rapid development and implementation of COVID-19 diagnostics has meant that normally stringent regulatory criteria have not been applied to all tests, with limited published data supporting assay performance in clinical settings. Here, in order to inform the deployment of PoCT in Australia, we compared the performance characteristics of five commercially available PoCT with (i) a commercially-available enzyme-linked immunosorbent assay (ELISA) and (ii) a recently described surrogate virus neutralisation assay, using samples from: (a) patients with RT-PCR-confirmed COVID-19; (b) patients who were RT-PCR negative but presented with respiratory symptoms during the peak of the pandemic in Australia, and (c) patients pre-COVID-19 pandemic.

## METHODS

### Clinical samples and patient populations

A testing panel was specifically developed to test PoCT devices for this study (Supplementary Dataset), consisting of three patient populations: (i) sera from 91 patients with SARS-CoV-2 detected by RT-PCR from upper and / or lower respiratory tract specimens; (ii) sera from 36 patients with seasonal coronavirus infections or other acute infections (e.g. dengue; CMV; EBV), and (iii) serum from a random cohort (56 patients) of the Australian population obtained in 2018.

To further assess specificity, one of the devices (Device 1, for which there was a surplus of tests) was tested against serum samples from 1,217 patients who were SARS-CoV-2 RT-PCR negative but presented to a hospital emergency department between 6^th^ February 2020 and 15^th^ April, spanning the initial peak of the COVID-19 pandemic in Australia.

Serum samples were obtained from a large academic hospital in Melbourne, Australia (Royal Melbourne Hospital, RMH), or the Victorian state reference laboratory for virology (Victorian Infectious Diseases Reference Laboratory, VIDRL). Convalescent patients were followed up at home by the RMH@Home Hospital in the Home (HITH) Team. Information on each cohort is provided in the Supplementary Dataset.

Cases were classified clinically as mild (not admitted to hospital), moderate (admitted to a hospital ward, but not the intensive care unit [ICU]) or severe (admitted to ICU). Of the 91 cases, 71 were mild, 17 were moderate and three were severe.

### RT-PCR

SARS-CoV-2 RNA was detected using the Coronavirus Typing assay (AusDiagnostics, Mascot, Australia), a two-step, hemi-nested multiplex tandem PCR, with seven coronavirus RNA targets plus a proprietary artificial sequence as an internal control. All positive samples underwent additional confirmatory testing for SARS-CoV-2 at VIDRL, using previously published primers (6).

### Serological PoCT

Serological PoCT devices were tested exactly as per the manufacturer’s stated instructions for use (IFU), including use of plastic droppers and buffers provided in the kits. Devices were provided through the Australian Government Therapeutic Goods Administration (TGA), based on their initial desktop evaluations of assay performance characteristics, and device availability at the time of the study (Supplementary Table 1). In brief, 10 ¼L of serum was added to the device, with addition of between 60 and 100 ¼L of the manufacturer’s provided buffer. Devices were incubated at room temperature according to the time period defined in the IFU (generally 10 to 15 minutes). All results were read as per the IFU. Testing was performed by laboratory technicians, all of whom had undergone competency training in the use of lateral flow assays. Testing of each sample in the serum panel was performed in duplicate, with a triplicate deciding test for discordant results. Any faint line present at test termination was considered a positive result. Results were recorded in a password-protected database available only to study investigators. All patient samples were de-identified.

### Enzyme-linked immunosorbent assay (ELISA)

ELISA testing was performed using a commercially available ELISA (Supplementary Table 1). The ELISA involves semi-quantitative detection of anti-SARS-CoV-2 IgA or IgG antibodies in serum through binding to a recombinant structural antigen (S1 domain of the Spike protein) fixed to reagent wells. If test sera contain anti-SARS-CoV-2 antibodies, a second incubation step using enzyme-labelled anti-human IgA or anti-human IgG will catalyse a colour reaction, detected by an optical density reader. Semiquantitative results were reported as a ratio as per the manufacturer’s IFU and interpreted as follows (i) ratio <0.8: negative result; (ii) ratio ≥ 0.8 to <1.1: borderline result, and (iii) ratio ≥ 1.1: positive result. Performance characteristics were determined using the same sera panel as the PoCT, along with 36 additional samples (33 samples from 19 patients with COVID-19 confirmed by RT-PCR; 2 samples from MERS-CoV positive patients and 1 serum from a SARS-CoV-1 positive patient) (Supplementary Dataset).

### SARS-CoV-2 Surrogate Virus Neutralization Test (sVNT)

To further assess antibody response, we used a recently described surrogate virus neutralization test (sVNT), that detects circulating neutralising antibodies in an isotype and species independent manner, based on antibody-mediated blockage of interaction between the ACE2 receptor protein and the receptor binding domain (RBD) of the viral spike protein (7). In brief, 10 ¼L of test serum is diluted with 90 ¼L of Sample Dilution Buffer and incubated with horseradish peroxidase conjugated SARSCoV-2 RBD protein (HRP-RBD), and the test solution is added to wells coated with fixed ACE2 receptors. The degree to which test serum inhibits binding of the HRP-RBD to ACE2 receptors, compared to control serum, is determined by optical density reading, with 20% inhibition and above considered a positive result. Sera tested in the sVNT included 169 samples from 110 patients with RT-PCR confirmed COVID-19 and 142 samples from 142 control patients; of which 36 samples were from patients with seasonal coronavirus or other acute infections, and 106 samples were from a random cohort of the Australian population obtained in 2016 and 2018 (Supplementary Dataset). A first round of testing on all samples followed the IFU; subsequently samples within the 10% coefficient of variation (CV) range as stated in the IFU (inhibition cut-off of 18–22%, n = 21), were repeated in duplicate to assess for inter-run variation.

### Statistical analysis

All statistical analyses were conducted using R (version 3.6.3) or GraphPad Prism (version 8.4.2). Binomial 95% confidence intervals (CI) were calculated for all proportions. Differences in non-normally distributed numerical data were calculated using the Wilcoxon Rank sum test. Receiver operating characteristic (ROC) area under the curve (AUC) analysis was performed in GraphPad Prism (version 8.4.2).

### Ethics

Ethical approval for this project was obtained from the Melbourne Health Human Research Ethics Committee (RMH HREC QA2020052).

## RESULTS

### Comparison of commercial ELISA vs. RT-PCR

The overall sensitivity for either IgA or IgG detection was 67.9% (95% CI 59.4–75.6%)) and specificity was 72.8% (95% CI 62.6–81.6%) (Table 1). The sensitivity for IgA or IgG detection increased to 93.8% (95% CI 85.0–98.3%) when only samples collected > 14 days post-symptom onset were considered (Table 2), and a significant rise in signal / cut-off ratio was observed for both IgA and IgG over time (P < 0.001) (Figure 1).

**Figure 1.**
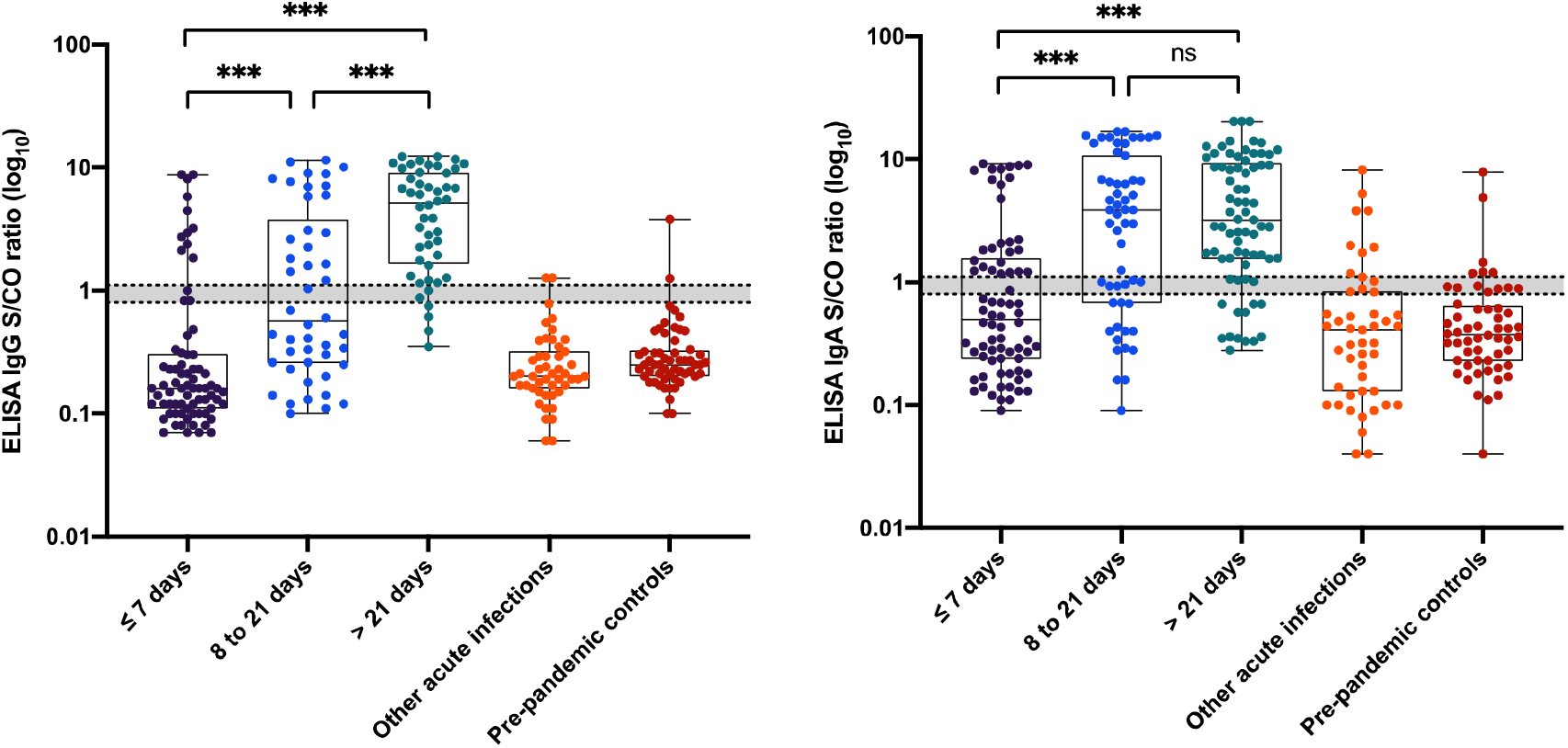
Distribution of signal / cut-off ratios obtained for the ELISA for (i) SARS-CoV-2 cases stratified by time post-symptom onset and (ii) control sera. Boxes represent median values and interquartile range, and whiskers represent maximum and minimum values. Dotted lines indicate the manufacturer’s cut-off values for interpretation of positive and negative test results, and the shaded grey area represents the range with borderline results. Abbreviations: *** indicates P value of < 0.0001; ns, not significant.

**Figure 2.**
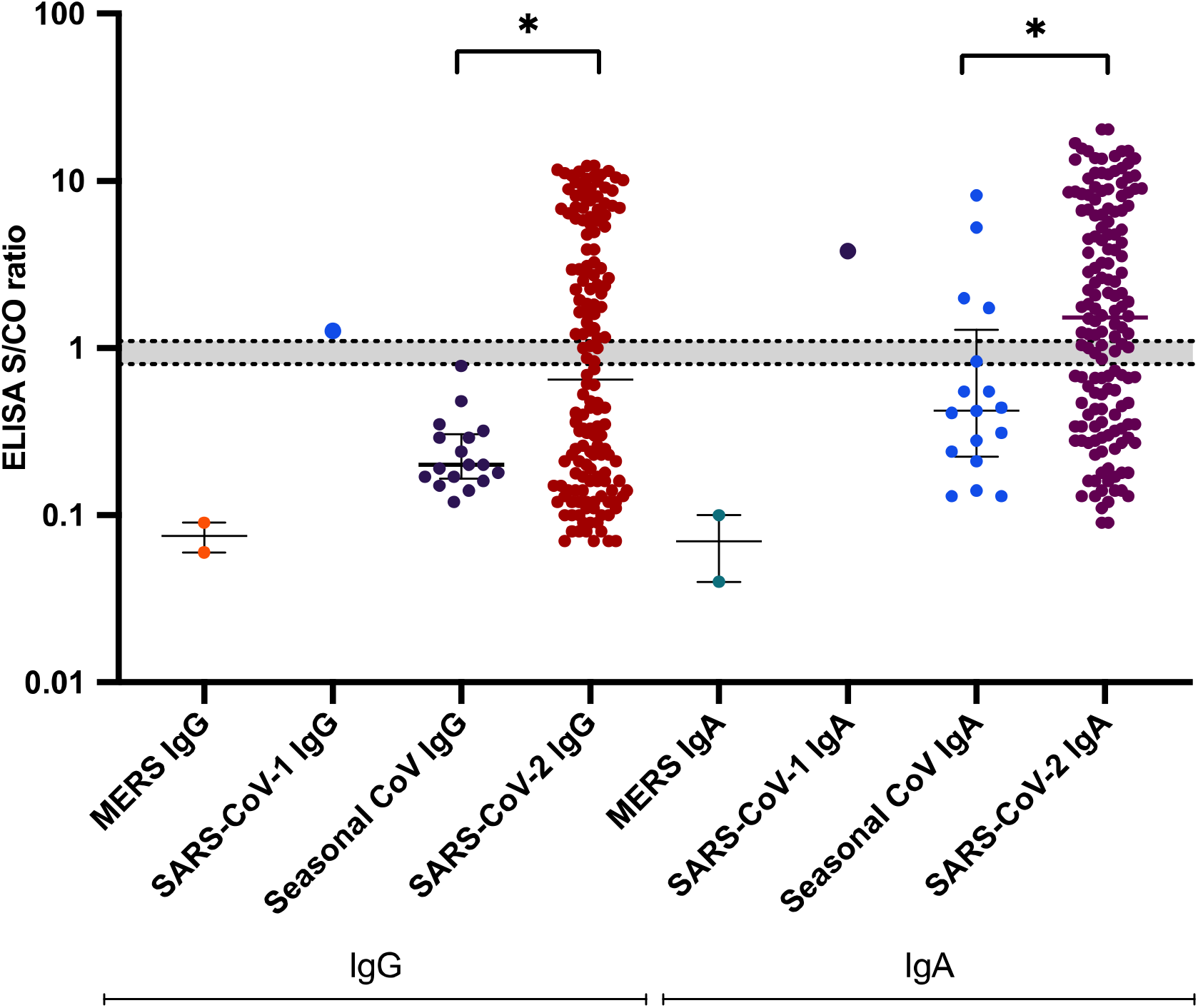
Distribution of signal / cut-off ratios obtained for the ELISA for SARS-CoV-2 cases and other human coronavirus infections. Lines represent median values and interquartile ranges. Abbreviations: * indicates P value of < 0.01.

**Table 1.** Comparative performance of serological assays with RT-PCR, at all sampling time points post-symptom onset.

| Performance Characteristic | Sensitivity (%) | Specificity (%) | Positive Predictive Value (%) | Negative Predictive Value (%) | Total (samples/ patients) |
| --- | --- | --- | --- | --- | --- |
| Test Assay | [95% CI] | [95% CI] | [95% CI] | [95% CI] |  |
| Device 1 IgM | 49.6 [41.0, 58.3] | 96.7 [90.8, 99.3] | 95.8 [88.1, 99.1] | 56.3 [48.2, 64.2] | 229/183 |
| Device 1 IgG | 46.7 [38.2, 55.4] | 98.9 [94.1, 99.97] | 98.5 [91.7, 99.96] | 55.5 [47.5, 63.2] | 229/183 |
| Device 1 IgM or IgG | **56.9 [48.2, 65.4]** | **95.6 [89.2, 98.8]** | **95.1 [88.0, 98.7]** | **59.9 [51.5, 67.9]** | 229/183 |
| Device 2 IgM | 51.8 [43.1, 60.4] | 97.8 [92.4, 99.7] | 97.3 [90.5, 99.7] | 57.6 [49.5, 65.6] | 229/183 |
| Device 2 IgG | 51.8 [43.1, 60.4] | 98.9 [94.1, 99.97] | 98.6 [92.5, 99.96] | 58.0 [49.8, 65.8] | 229/183 |
| Device 2 IgM or IgG | **51.8 [43.1, 60.4]** | **97.8 [92.4, 99.7]** | **97.3 [90.5, 99.7]** | **57.7 [49.6, 65.6]** | 229/183 |
| Device 3 IgM | 13.1 [8.0, 20.0] | 96.7 [90.8, 99.3] | 85.7 [63.7, 97.0] | 42.8 [36.0, 49.8] | 229/183 |
| Device 3 IgG | 59.9 [51.1, 68.1] | 100 [96.1, 100] | 100 [95.6, 100] | 62.6 [54.2, 70.4] | 229/183 |
| Device 3 IgM or IgG | **60.6 [51.9, 68.8]** | **96.7 [90.8, 99.3]** | **96.5 [90.1, 99.3]** | **62.2 [53.8, 70.2]** | 229/183 |
| Device 4<sup>#</sup> | **68.6 [60.1, 76.3]** | **97.8 [92.4, 99.7]** | **97.9 [92.7, 99.8]** | **67.7 [59.0, 75.5]** | 229/183 |
| Device 5 IgM | **39.0 [30.7, 47.7]** | **100 [96.1-100]** | **100 [93.3, 100]** | **52.6 [44.9, 60.2]** | 228/182 |
| Device 5 IgG | **58.8 [50.7, 67.2]** | **100 [96.1-100]** | **100 [95.6, 100]** | **62.2 [53.8, 70.0]** | 228/182 |
| Device 5 IgM or IgG | **61.0 [52.3, 69.3]** | **100 [96.1-100]** | **100 [95.7, 100]** | **63.4 [55.1, 71.3]** | 228/182 |
| ELISA IgA | 65.7 [57.1, 73.6] | 73.9 [63.7, 82.5] | 78.9 [70.3, 86.0] | 59.1 [49.6, 68.2] | 229/183 |
| ELISA IgG | 56.2 [47.5, 64.7] | 97.8 [92.4, 99.7] | 97.5 [91.2, 99.7] | 60.0 [51.7, 67.9] | 229/183 |
| ELISA IgA or IgG | **67.9 [59.4, 75.6]** | **72.8 [62.6, 81.6]** | **78.8 [70.3, 85.8]** | **60.4 [50.6, 69.5]** | 229/183 |
| sVNT at 20% inhibition cut-off | **62.7 [55.0, 70.0]** | **94.4 [89.2, 97.5]** | **93.0 [86.6, 96.9]** | **68.0 [61.0, 74.5]** | 311/252 |
| sVNT at 25% inhibition cut-off | 60.9 [53.2, 68.4] | 99.3 [96.1, >99.9] | 99.0 [94.8, >99.9] | 68.1 [61.3, 74.4] | 311/252 |
| sVNT at 30% inhibition cut-off | 55.6 [47.8, 63.3] | 100 [97.4, 100] | 100 [96.2, 100] | 65.4 [58.7, 71.8] | 311/252 |
| sVNT at 20% inhibition cut-off, including equivocal range (18-22% inhibition) | 61.5 [53.8, 68.9] | 99.3 [96.1, >99.9] | 99.0 [94.8, >99.9] | 68.4 [61.6, 74.7] | 311/252 |

<sup>#</sup>Single test line captures IgM and IgG antibodies

**Table 2.** Comparative performance of serological assays with RT-PCR for samples collected > 14 days post symptom onset.

| Performance Characteristic | Sensitivity (%) | Specificity (%) | Positive Predictive Value (%) | Negative Predictive Value (%) | Total (samples/patients) |
| --- | --- | --- | --- | --- | --- |
| Test Assay | [95% CI] | [95% CI] | [95% CI] | [95% CI] |  |
| Device 1 IgM | 69.2 [56.6, 80.1] | 96.7 [90.8, 99.3] | 93.8 [82.8, 98.7] | 81.7 [73.1, 88.4] | 157/155 |
| Device 1 IgG | 80.0 [68.2, 88.9] | 98.9 [94.1, 99.97] | 98.1 [89.9, 99.95] | 87.5 [79.6, 93.2] | 157/155 |
| Device 1 IgM or IgG | **84.6 [73.5, 92.4]** | **95.6 [89.2, 98.8]** | **93.2 [83.5, 98.1]** | **89.8 [82.0, 95.0]** | 157/155 |
| Device 2 IgM | 78.5 [66.5, 87.7] | 97.8 [92.4, 99.7] | 96.2 [87.0, 99.5] | 86.5 [78.5, 92.4] | 157/155 |
| Device 2 IgG | 78.5 [66.5, 87.7] | 98.9 [94.1, 99.97] | 98.1 [89.9, 99.95] | 86.7 [78.6, 92.5] | 157/155 |
| Device 2 IgM or IgG | **78.5 [66.5, 87.7]** | **97.8 [92.4, 99.7]** | **96.2 [87.0, 99.5]** | **86.5 [78.5, 92.4]** | 157/155 |
| Device 3 IgM | 10.8 [4.4, 20.9] | 96.7 [90.8, 99.3] | 70 [34.8, 93.3] | 60.5 [52.5, 68.5] | 157/155 |
| Device 3 IgG | 90.8 [81.0, 96.5] | 100 [96.1, 100] | 100 [93.9, 100] | 93.9 [87.2, 97.7] | 157/155 |
| Device 3 IgM or IgG | **90.8 [85.1, 96.5]** | **96.7 [90.8, 99.3]** | **95.2 [86.5, 99.0]** | **93.6 [86.8, 97.7]** | 157/155 |
| Device 4<sup>#</sup> | **93.8 [85.0, 98.3]** | **97.8 [92.4, 99.7]** | **96.8 [89.0, 99.6]** | **95.7 [89.5, 98.7]** | 157/155 |
| Device 5 IgM | **59.4 [46.4, 71.5]** | **100 [96.1, 100]** | **100 [90.8, 100]** | **77.3 [68.7, 84.5]** | 156/154 |
| Device 5 IgG | **93.8 [85.0, 98.3]** | **100 [96.1, 100]** | **100 [94.1, 100]** | **95.8 [89.7, 98.9]** | 156/154 |
| Device 5 IgM or IgG | **93.8 [84.8, 98.3]** | **100 [96.1, 100]** | **100 [94.1, 100]** | **95.8 [89.7, 98.9]** | 156/154 |
| ELISA IgA | 89.2 [79.1, 95.6] | 73.9 [63.7, 82.5] | 70.7 [59.7, 80.3] | 90.7 [81.7, 96.2] | 157/155 |
| ELISA IgG | 92.3 [83.0, 97.5] | 97.8 [92.4, 99.7] | 96.8 [88.8, 99.6] | 94.7 [88.1, 98.3] | 157/155 |
| ELISA IgA or IgG | **93.8 [85.0, 98.3]** | **72.8 [62.6, 81.6]** | **70.9 [60.1, 80.2]** | **94.4 [86.2, 98.4]** | 157/155 |
| sVNT at 20% inhibition cut-off | **91.2 [81.8, 96.7]** | **94.4 [89.2, 97.5]** | **88.6 [78.7, 94.9]** | **95.7 [90.9, 98.4]** | 210/205 |
| sVNT at 25% inhibition cut-off | 89.7 [79.9, 95.8] | 99.3 [96.1, >99.9] | 98.4 [91.3, >99.9] | 95.3 [90.5, 98.1] | 210/205 |
| sVNT at 30% inhibition cut-off | 88.2 [78.1, 94.8] | 100 [97.4, 100] | 100 [94.0, 100] | 94.7 [89.8, 97.7] | 210/205 |
| sVNT at 20% inhibition cut-off, including equivocal range (18-22% inhibition) | 91.2 [81.8, 96.7] | 99.3 [96.1, >99.9] | 98.4 [91.5, >99.9] | 95.9 [91.3, 98.5] | 210/205 |

<sup>#</sup>Single test line captures IgM and IgG antibodies

ROC AUC analysis was performed for both IgA and IgG. Overall, the IgA ROC AUC was 0.74 (95% CI 0.69–0.81) and the IgG was 0.66 (95% CI 0.59–0.72) (Supplementary Figures 1A and 1B). However, when only samples collected > 14 days post-symptom onset were included, the ROC AUC increased to 0.92 (95% CI 0.87–0.96) for IgA and 0.97 (95% CI 0.94–0.99) for IgG (Supplementary Figures 1C and 1D). No cross-reactivity with seasonal coronavirus infection was observed for IgG, although 4/17 (23.5%) samples from patients with seasonal coronavirus (two HKU1, one NL63 and one OC43) were positive for IgA. Neither of the two samples with anti-MERS-CoV antibodies displayed cross-reactivity for SARS-CoV-2 IgA or IgG, but one sample with anti-SARS-CoV-1 antibodies had positive results for SARS-CoV-2 IgA and IgG (ratios 3.81 and 1.26, respectively) (Figure 3).

**Figure 3.**
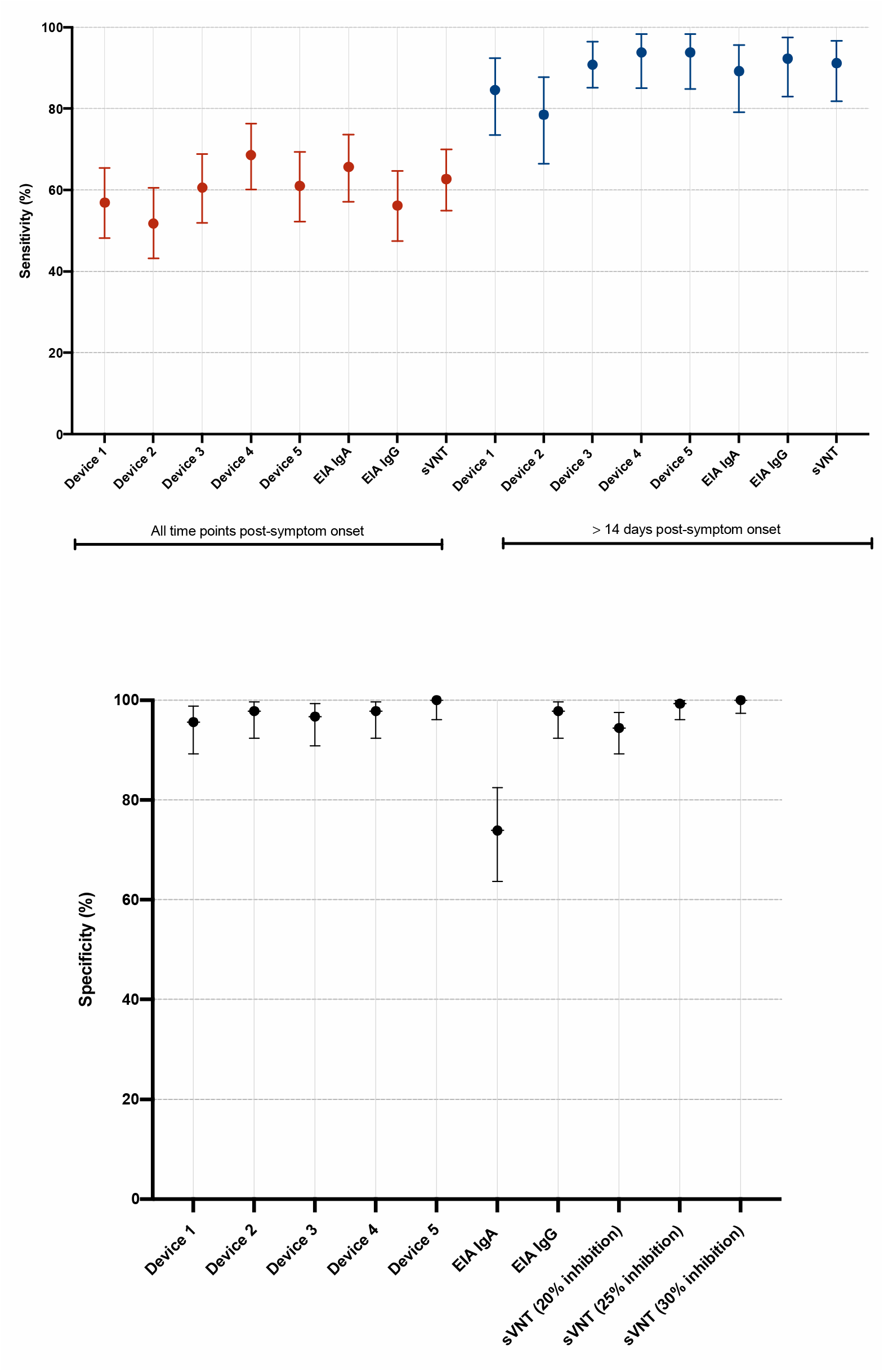
(A) Sensitivity and (B) specificity of five different serological point-of-care devices and one commercial enzyme immunoassay compared to reverse-transcriptase SARS-CoV-2 PCR (RTPCR).

### Comparison of PoCT and RT-PCR

We compared the sensitivity and specificity of five PoCT devices, using RT-PCR as our reference standard, and interpreting PoCT results as positive when either an IgM or IgG result was read as positive. Overall, the sensitivities ranged from 51.8% (95% CI 43.1 to 60.4%) to 68.6% (95% CI 60.1–76.3%), and specificities from 95.6% (95% CI 89.2–98.8%) to 100.0% (95% CI 96.1–100.0%) (Table 1 and Figures 3A and 3B). When only samples collected > 14 days were considered, the sensitivities ranged from 78.5% (95% CI 66.5–87.7%) to 93.8% (95% CI 85.0–98.3%) (Table 2).

Using Device 1 (for which there was a surplus of kits), additional testing was conducted on 1,217 samples from patients who presented with respiratory symptoms but tested RT-PCR negative for SARS-CoV-2. In total, 39/1,217 (3.2%) samples tested positive for IgM and /or IgG. On further testing, 6/39 samples (15.4%) tested positive to IgA and / or IgG using the ELISA assay, of which one was confirmed by sVNT (inhibition 63.9%) when an inhibition cut-off of 20% was employed (see below). This patient presented 21 days following symptom onset, with significant epidemiological risk factors for SARS-CoV-2 acquisition, and likely represents a true infection.

Using the highest performance device characteristics (sensitivity 68.6% [Device 4] and specificity > 99.9% [Device 5]) and lowest performance characteristics (sensitivity 51.8% [Device 2] and specificity 95.6% [Device 1]) as hypothetical ‘best’ and ‘worse’ scenarios respectively, the performance of PoCT was assessed across a range of SARS-CoV-2 population prevalence estimates(0.1%, 1%, 5% and 10%). (Table 3 and Supplementary Figure 1). With the best performing PoCT characteristics at an estimated SARS-CoV-2 period prevalence in Australia of 0.03%, the positive predictive value was only 17.1%.

**Table 3.** Performance characteristics of ‘best case’ and ‘worst case’ point of care devices across a range of population prevalence estimates.

| Device characteristics | SARS-CoV-2 prevalence (%) |  |  |  |
| --- | --- | --- | --- | --- |
|  | 0.1 | 1 | 5 | 10 |
| **Best case <sup>#</sup>** |  |  |  |  |
| PPV (%) | 40.7 | 87.4 | 97.3 | 98.7 |
| NPV (%) | 99.9 | 99.7 | 98.4 | 96.6 |
| FP/1000 tests | 1 | 1 | 1 | 0.9 |
| FN/1000 tests | 0.3 | 3.1 | 15.7 | 31.4 |
| **Worst case <sup>\*</sup>** |  |  |  |  |
| PPV (%) | 1.2 | 10.6 | 38.2 | 56.7 |
| NPV (%) | 99.9 | 99.5 | 97.4 | 94.7 |
| FP/1000 tests | 44 | 43.6 | 41.8 | 39.6 |
| FN/1000 tests | 0.5 | 4.8 | 24.1 | 48.2 |

### Comparison of sVNT and RT-PCR

In total, 311 samples were also tested using the sVNT assay. Applying a 20% inhibition cut-off and using RT-PCR as the reference standard, the sensitivity of sVNT was 62.7% (95% CI 55.0–70.0%); this increased to 91.2%% (95% CI 81.8–96.7%) when only samples collected > 14 days post-symptom onset were considered (Tables 1 and 2). Specificity was 94.4% (95%CI 89.2–97.5%), with cross-reaction observed for eight samples (Figure 4 and Supplementary Table 2). Increasing the inhibition cut-off to 25%, or repeating samples with an initial inhibition score between 18–22% improved the specificity to 99.3% (95% CI 96.1–99.9%) with little change in sensitivity (Tables 1 and 2). The coefficient of variation (% CV) for the in-house control sample with respect to the percentage inhibition was 10.8% between runs, and 5.8% within-run.

**Figure 4.**
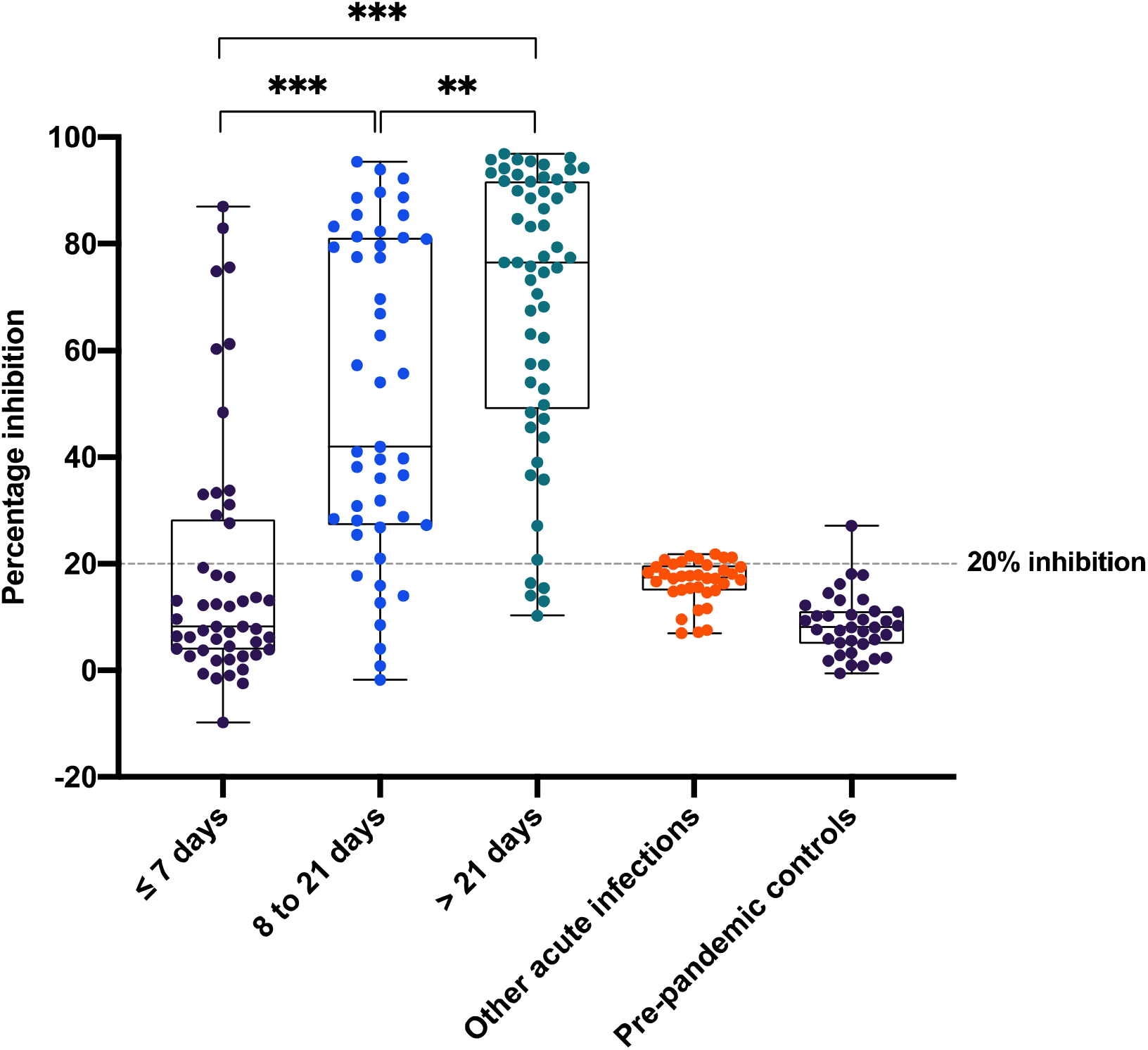
Distribution of surrogate virus neutralization testing (sVNT) percentage inhibition for SARS-CoV-2 cases stratified by time post-symptom onset and control sera. Boxes represent median values and interquartile range, and whiskers represent maximum and minimum values. Abbreviations: *** indicates P value of < 0.0001; ***indicates P value of <0.01; ns, not significant.

## DISCUSSION

Accurate laboratory testing is integral to the prevention and control of COVID-19. The unprecedented scale of diagnostic testing has led to the rapid development and implementation of a large range of diagnostic assays for SARS-CoV-2, including serological tests. However, there are limited peer-reviewed data on the performance characteristics of serological tests, and in order to best inform the implementation of these assays, high-quality post-market validation data are urgently needed to guide laboratories, public health agencies and governments in the appropriate and responsible use of such tests (8).

In this study, we assessed the performance characteristics of five serological PoCT, one commercial ELISA and one commercial novel surrogate virus neutralisation test against a large serum panel from a cohort of over 100 patients with RT-PCR confirmed SARS-CoV-2. In keeping with other studies (9), the sensitivity of all assays was low (< 70%) when all sample collection time points were considered. However, as expected given the reported antibody response to SARS-CoV-2 infection, sensitivity increased considerably when samples collected > 14 days post-symptom onset were assessed (10), with the majority achieving sensitivities over 90%. Our findings provide further support for recent commentary suggesting that current serological assays have limited, if any, role in the diagnosis of acute COVID-19, with RT-PCR remaining the gold standard for diagnosis in the acute setting (11, 12). Specificities for PoCT ranged from 92.4%-100%; it is possible this may reflect differences in the antigens used in each assay, although specific information about the SARS-CoV-2 recombinant antigen used in the assay was not described for most PoCT. In keeping with previous reports (13, 14), when both IgA and IgG components of the ELISA were considered, specificity was low (72.8%), but considering IgG alone, specificity increased to 97.8%.

This study is one of the first to utilise a recently described sVNT assay (7). Previous work describing the development of this assay reported a 95–100% sensitivity and 100% specificity using cohorts in Singapore and China (7). In our cohort at > 14 days post symptom onset we achieved sensitivity of

91.2% (95% CI 81.8–96.7) and specificity of 94.4% (95%CI 89.2–97.5%). Although limited clinical data are available on the cohort used to develop and validate the assay (7), it is possible that our relatively mild clinical cohort may generate lower antibody titres than a more severely unwell cohort, potentially influencing sensitivity of the assay (10, 15). Of note, the majority of our non-specific (false positive) samples in the sVNT assay recorded inhibition just over the 20% cut-off. However, specificity improved when either: (i) a higher inhibition cut-off was used, or (ii) samples within an arbitrary range (based on the IFU reported %CV of the assay) were tested in triplicate. It is possible that populations of different ethnic background or geographic specific cut-offs may be required for the sVNT assay as suggested by the kit provider; in our low-prevalence setting where the test is more likely to act as a confirmatory assay, raising the inhibition cut-off to 25% increased the specificity to 99.3% (95% CI 96.1–99.9%), thus improving clinical utility. Alternatively, introduction of an equivocal range for the assay with repeat testing for samples within this range, would be another approach to mitigate potential assay variation.

In contrast to acute diagnosis, there are settings where high-quality serological assays will have utility, including (i) defining antibody prevalence in key populations such as frontline workers; (ii) determining the extent of COVID-19 infection within the community; (iii) identifying individuals for further evaluation of therapeutic immunoglobulin donation, and (iv) vaccine development and evaluation. For (iii) and (iv), it is essential to have a good quantification of the functional neutralizing antibodies among donors or vaccines. However, in order to appropriately deploy serological testing, it is critical to understand the limitations of test performance in the epidemiological context in which tests are used. This is particularly important in a setting such as Australia, which, based on the number of reported cases of COVID-19 (7,060 cases as of 18th May, 2020), has an estimated COVID-19 period prevalence of 0.03% (16). As such, even with highly sensitive and specific serological tests, the majority of positive results are likely to represent false positives. When considering the use of serology to inform policies relating to relaxing of physical distancing interventions, specificity of the assay becomes critical. If the majority of those identified as immune are actually false positive results, then the threshold to maintain immunity within the community will not be achieved (17).

Analogous to HIV testing in low-prevalence settings (18), serological testing for SARS-CoV-2 may require a ‘two-step’ approach, whereby a sensitive high-throughput ‘screening’ assay is followed by a high specificity assay for confirmation (e.g. neutralisation testing or Western blot). This approach could facilitate seroepidemiological studies in low-prevalence settings, which are required to better understand the extent of COVID-19 infection at a population-level. Ongoing questions remain however, about the duration and type of antibody response to SARS-CoV-2, particularly around the protective effect of neutralising antibodies against future re-infection (10). Accordingly, the concept of an ‘immunity passport’ that facilitates return to workplaces or school should be interpreted with caution, and the World Health Organization (WHO) currently recommends the use of PoCT immunodiagnostic assays in research settings only, and not for clinical decision making until further evidence is available (19).

A key strength of this study was our systematic collection of convalescent samples. By establishing a community collection platform, we tested over 50 patients who were more than 21 days post-symptom onset. Ideally, validation of serological assays should be performed against a testing panel that includes samples from: (i) patients at acute and convalescent stages of infection (to assess sensitivity), and (ii) patients with other human coronavirus infections (to assess specificity). Given the range of serological assays now available, there is a critical need for standardised protocols, including reference standards, across laboratories when conducting evaluations of emerging serological assays.

In summary, our data describe the performance characteristics of five PoCT devices, a commercially available ELISA assay and a newly developed surrogate virus neutralisation test. Overall, our findings are in keeping with recent position statements that note that serological assays have limited, if any, role in the diagnosis of acute COVID-19 infection. Our data strongly suggest that current PoCT devices should not be used in the diagnosis of acute COVID-19, or as the sole assay in population- level serosurveys. . Nevertheless, there are settings where high-quality serological assays will have clinical utility. The curated panel of samples assembled for this study is being expanded and provides a valuable repository for rapid validation of new serological assays as they become available.

## Data Availability

We have included all relevant data.

## ACKNOWLEDGEMENTS

We thank the scientific staff of the Pathology Department at the Royal Melbourne Hospital, particularly Maria Bisignano, Siobhan Carne, Elizabeth Shilling and Merina Gurung. We also thank the Serology Department of the Victorian Infectious Diseases Reference Laboratory (VIDRL), Francesca Mordant and Kanta Subbarao from the Department of Microbiology and Immunology, University of Melbourne. Additionally, we thank the RMH@Home staff, including Beverley Cox and Samuel Strapps and the staff of the RMH Emergency Department, particularly Dr. Mark Putland. We especially thank the patients who contributed to this study. We thank Professor Linfa Wang and his team at Duke-NUS Medical School for providing the sVNT kit to us before it is commercially available.

## FUNDING

DAW is supported by an Investigator Grant from the National Health and Medical Research Council (NHMRC) of Australia (APP1174555). BPH is supported by a NHMRC Practitioner Fellowship (APP1105905). KB is supported by an NHMRC Postgraduate Scholarship (GNT1191321). This work was supported by a grant from the NHMRC Medical Research Future Fund (APP2002317).

## CONFLICTS

All authors: no conflicts.

## Notes

### Competing Interest Statement

The authors have declared no competing interest.

